# Tuberculosis-associated hemophagocytic lymphohistiocytosis: diagnostic challenges and determinants of outcome

**DOI:** 10.1101/2023.11.14.23298501

**Authors:** Lisa Kurver, Timothy Seers, Suzanne van Dorp, Reinout van Crevel, Gabriele Pollara, Arjan van Laarhoven

## Abstract

**Background:** Tuberculosis (TB) can induce secondary hemophagocytic lymphohistiocytosis (HLH), a severe inflammatory syndrome with high mortality. To improve insight into optimal diagnostic and treatment strategies, we integrated all published reports of adult HIV-negative TB-associated HLH (TB-HLH) globally to define clinical characteristics and therapeutic approaches associated with improved survival.

**Methods:** PubMed, Embase, and Global Index Medicus were searched for eligible records. TB-HLH cases were categorized into patients with a confirmed TB diagnosis receiving antituberculosis treatment while developing HLH, and patients presenting with HLH of unknown cause later diagnosed with TB. We integrated patients’ clinical characteristics, diagnostic test results, and pre-specified parameters associated with survival into a logistic regression model.

**Results:** We identified 115 individually reported cases, 45 (39.1%) from low TB incidence countries (<10/100.000 per year). Compared to HLH patients with known TB (n=21), patients with HLH of unknown cause (n=94), more often had extrapulmonary TB (88.3% vs. 66.7%), while the opposite was true for pulmonary disease (59.6% vs. 91.5%). Overall, *Mycobacterium tuberculosis* was identified in the bone marrow in 78.4% of patients for whom examination was reported (n=74). Only 10.5% (4/38) of patients tested had a positive tuberculin skin test or interferon gamma release assay. In-hospital survival was 71.9% (69/96) in those treated for TB and 0% (18/18) in those who did not receive antituberculosis treatment (p < 0.001).

**Conclusions:** Tuberculosis should be considered as a cause of unexplained HLH. TB-HLH is probably under-reported, and the diagnostic work-up of HLH patients should include bone marrow examination for evidence of *M. tuberculosis* infection. Prompt initiation of antituberculosis treatment will likely improve survival.

**Key points:** Hemophagocytic lymphohistiocytosis is an underreported complication of tuberculosis, often manifesting as extrapulmonary or miliary disease. TST and IGRA mostly show an anergic response. Threshold should be low bone marrow investigation for evidence of *M. tuberculosis,* and commencement of antituberculosis treatment.

## Introduction

Hemophagocytic lymphohistiocytosis (HLH) is a severe hyperinflammatory syndrome characterized by elevated pro-inflammatory cytokine production, excessive macrophage activation, and phagocytosis of mature blood elements [1,2]. Contrary to primary HLH, which is driven by known genetic mutations, secondary HLH is caused by malignancies, auto-immune and auto-inflammatory disorders (then termed macrophage activation syndrome), or infections, including tuberculosis (TB) [3]. Overall, HLH is associated with high mortality [4]. Immunomodulation with corticosteroids, IL-1 receptor antagonism or T-cell directed chemotherapy may be necessary to curb inflammation. However, in the case of secondary HLH of infectious origin, pathogen-directed treatment is crucial [5,6].

Diagnosis of TB disease relies on microbiological identification of *Mycobacterium tuberculosis* (*M. tuberculosis*) by culture, microscopy, or molecular testing, and can be supported by histopathological evidence of granulomatous inflammation. Low suspicion for TB may preclude use of appropriate TB diagnostics in low TB-endemic settings, while in high TB-endemic settings, diagnosing HLH may be difficult if bone marrow examination and other tests for HLH are unavailable. T-cell-based assays for immune memory to *M. tuberculosis*, such as the tuberculin skin test (TST) or interferon gamma release assay (IGRA) are sometimes used to exclude TB, but their diagnostic accuracy might be affected by the concurrent immune dysregulation associated with HLH.

Currently, understanding of optimal diagnostic strategies and therapeutic approaches in TB-HLH is limited because data is largely confined to case reports and small case series. A recent review on clinical characteristics of TB-HLH included a heterogenous patient population including children and lacked pre-defined patient categorization or multivariate assessment of the determinates of outcomes [7]. This limits the ability to draw firm conclusions on how to best identify patients with TB-HLH, the investigations with greatest diagnostic yield, and the interventions that most impact survival. To address these unanswered research questions, we undertook a systematic review of all TB-HLH cases described to date according to PRISMA guidelines performing a patient-level meta-analysis of all reported clinical parameters. We specified two separate categories of TB-HLH patients: those with a confirmed TB diagnosis already receiving antituberculosis treatment at the time of HLH diagnosis, and patients presenting with HLH of unknown etiology, in whom TB was later diagnosed. The standardized collection of information from the reported TB-HLH cases allowed us to integrate findings on the clinical presentation, diagnosis, treatment, and outcomes in TB-HLH.

## Methods

### Search strategy and selection criteria

We defined and followed a pre-specified strategy according to PRISMA guidelines [8] registered with the international prospective register of systematic reviews (PROSPERO, CRD42022349077 [9]). We searched PubMed, Embase, and Global Index Medicus for records using “Tuberculosis” AND “Hemophagocytic lymphohistiocytosis” OR “Macrophage activation syndrome” and synonyms and abbreviations, indexed before October 23^rd^ 2022, without language or publication type restrictions in our Search Strategy (**Supplementary table 1**). Duplicate records were removed using Endnote and Rayyan.[10] First, one reviewer (LK) screened all titles and abstracts using the exclusion criteria: 1) the described patient was not diagnosed with TB; 2) the record contained no primary data of at least one patient; or 3) all described patients were HIV co-infected, as in-between patient differences in immune status (CD4+ T cell count) would cause more heterogeneity and create an overlap with HIV-associated immune reconstitution inflammatory syndrome. Next, two reviewers (GP and AvL) screened the remaining titles and abstracts again using the inclusion criteria. Records were included if they described at least one patient meeting both of the following criteria: 1) patient with tuberculosis and 2) hemophagocytic lymphohistiocytosis. Disagreements were resolved through discussion and mutual agreement by all three reviewers.

### Data extraction, eligibility and bias assessment

Full-text records were then assessed for eligibility by one reviewer (LK or TS) during data extraction using a standardized form. For each record, we collected data relating to 1) record characteristics; 2) patient clinical parameters; 3) TB disease features and therapy; 4) HLH characteristics and therapies; 5) TST / IGRA test results; and 6) patient outcomes. A detailed description of the collected data is provided in **Supplementary Table 2**. We assessed the risk of bias using the Joanna Briggs Institute critical appraisal tool for case reports [11]. We assessed the risk of bias on eight domains, including: 1) demographic characteristics; 2) medical history; 3) clinical condition; 4) diagnostics; 5) intervention and treatment; 6) post-intervention clinical condition; 7) adverse events; 8) takeaway lesson. Reports with missing data on any one the following key variables were excluded by the first reviewer: sex; age; tuberculosis localization; use of antituberculosis treatment at the onset of HLH; and outcome. A second reviewer (one of SvD, RvC, GP, AvL) reviewed all coded data for accuracy, so that all eligible studies received independent assessment from two reviewers. Disagreements were resolved through discussion and mutual agreement by both reviewers for each record in question.

Missing blood biochemical test results (continuous variables) were coded as missing. Co-morbidities were assumed to be absent if not reported, except for *M. tuberculosis* bone marrow examination which was scored as ‘present’ when either microbiological (culture, staining for acid-fast bacilli, or PCR) or histological evidence of *M. tuberculosis* were reported, ‘absent’ when the absence of microbiological or histological evidence of *M. tuberculosis* was reported and otherwise as ‘not tested’, as we did not assume that all bone marrow examinations included *M. tuberculosis*-diagnostics. Continuous values were converted to standard units when necessary.

### Case definitions and classifications

Identified patients were categorized into two groups: patients with known TB developing HLH as a paradoxical reaction while receiving antituberculosis treatment; and patients presenting with HLH of unknown cause who were later diagnosed with TB, and who therefore were not receiving antituberculosis treatment at the onset of HLH.

### Data analysis and visualization

Data were analyzed using SPSS 27th edition [12]. Chi-square tests (nominal variables) and Mann-Whitney U tests (continuous variables) were used to assess differences between groups. To determine associations with TB-HLH mortality, we built a logistic regression model. First, we explored association with mortality for sex, variables reported to influence HLH mortality (age, presence of comorbidities, hepatomegaly, hemoglobin and ferritin levels) [13–15], as well as steroid therapy [16], supplemented by covariates relevant to TB-HLH: TB localization, evidence of *M. tuberculosis* in the bone marrow and receiving antituberculosis treatment during the disease episode. Covariates univariately associated with survival were tested in a multivariable regression model. Data were visualized using GraphPad Prism version 10.0.2 [17] and R 4.1.3 [18] using the R packages “xlsx” [19], “maps” [20], and “ggplot2” [21].

## Results

We identified 641 records from the database search (**Figure 1**). After deduplication, screening of titles and abstracts (417 records), and full text screening (202 records), 100 publications were included. An additional 6 records that had not been picked up by our search criteria were identified via manual reference searches of the same databases. As missing data for our key variables was an exclusion criterion, the risk of bias was uniformly assessed as low **(Supplementary Table 3).** The 106 included records described 115 patients, reported from 34 countries across all continents except Antarctica (**Figure 2**). Reports from the 20 WHO-defined high TB-burden countries for 2021-2025 comprising 84% of the global TB burden [22], only comprised 35.6% (41/115) of cases. Reports from low TB-incidence (< 10 / 100.000 [23]) comprised 39.1% (45/115) of cases, with only one third (15/45) of these patients originating from a moderate to high incidence country (≥ 10 / 100.000).

**Figure 1:**
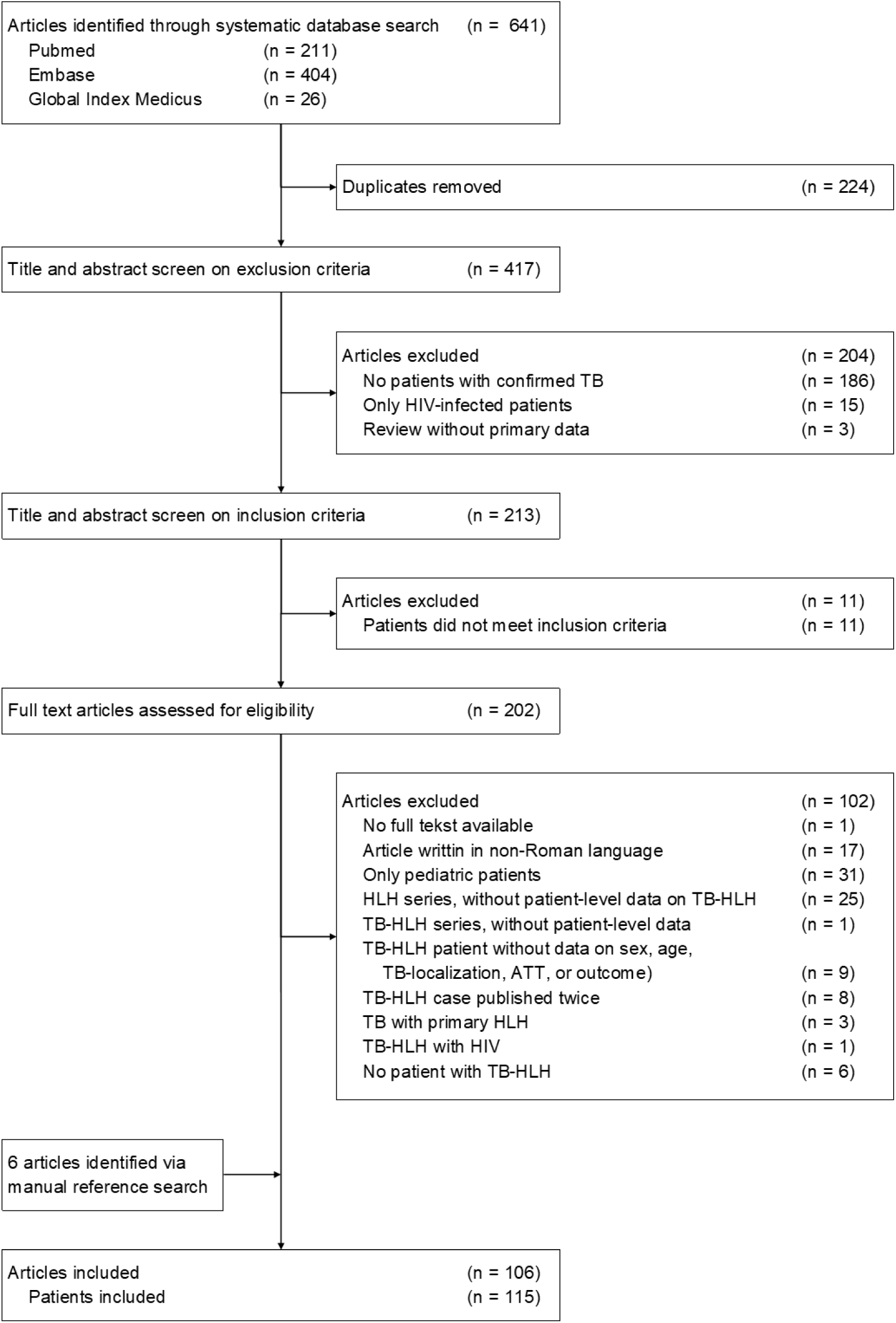
PRISMA diagram of articles reporting one or more cases of TB-HLH. **Abbreviations: TB** tuberculosis; **TB-HLH** tuberculosis associated hemophagocytic lymphohistiocytosis; **ATT** antituberculosis treatment.

**Figure 2:**
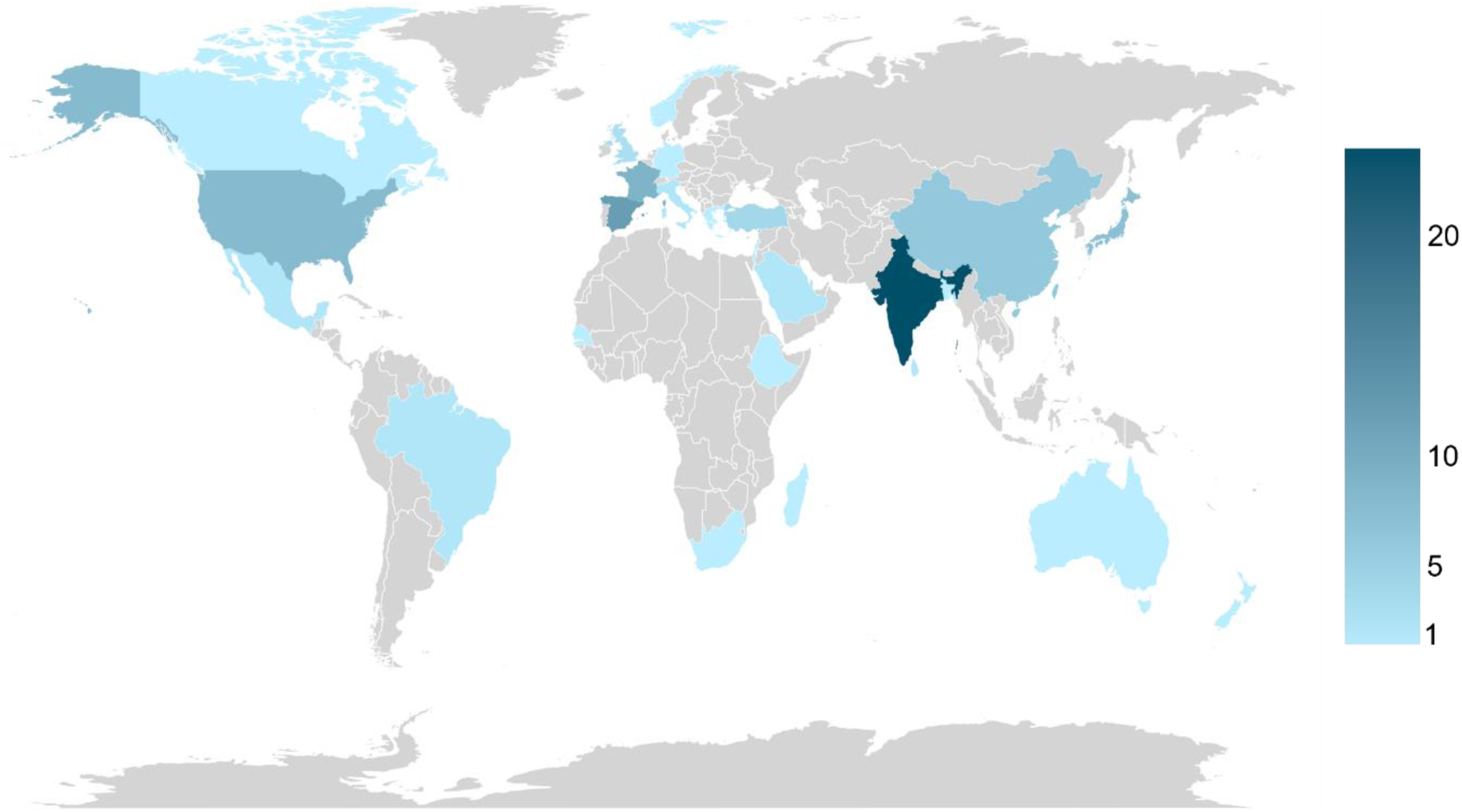
Geographical distribution of TB-HLH cases: number of reported TB-HLH cases according to country of origin for published report. Grey: no TB-HLH cases reported. **Abbreviations: TB-HLH** tuberculosis associated hemophagocytic lymphohistiocytosis.

Patients were mostly male (69.6%) with a median age of 43 (IQR 30-63), and approximately half had one or more comorbidities (**Table 1**). HLH-criteria were incompletely assessed, with limited availability of NK-cell activity and soluble IL-2 receptor (recorded in 5.2% and 11.3% of cases respectively). Consequently, out of the 8 HLH-2004 criteria for secondary HLH, a median of 5 (IQR 4-6) were assessed, and 66.1% cases fulfilled ≥5 criteria required to make a diagnosis of HLH (**Supplementary Figure 1**). The minority of TB-HLH cases developed HLH while already receiving antituberculosis treatment for a confirmed TB diagnosis (n=21) a median of 14 days (IQR 7-36 days) after start antituberculosis treatment. Most TB-HLH cases initially presented with HLH of unknown etiology and a TB diagnosis was made later (n=94). These two groups were similar in sex, age, presence of fever, splenomegaly, presence of hemophagocytosis in bone marrow, spleen, and/or lymphnodes, hyperferritinemia and other HLH-criteria, as well as underlying comorbidities, except for renal disease (**Table 1, Supplementary Table 4**). Notably, the diagnosis of TB-HLH could not be ruled out based on ferritin alone, as hyperferritinemia (≥ 500 µg/L, HLH-2004 cut-off [16]) was reported in 71.3%, and ferritin levels ≥ 2000 µg/L (H-score minimum [24]) were only reported in 38.3%.

**Table 1:** Patient characteristics and disease characteristics at the time of HLH presentation.

|  | On ATT at HLH<br>onset<br>N = 21 | Not on ATT at HLH<br>onset<br>N = 94 | p-value |
| --- | --- | --- | --- |
| **Patient characteristics** |  |  |  |
| Female, n (%) | 6 (28.6) | 29 (30.9) | 0.84 |
| Age, y, median (IQR) | 37.0 (28.5-59.0) | 45.0 (30.8-63.0) | 0.66 |
| Comorbidities, n (%) | 9 (42.9) | 49 (52.1) | 0.44 |
| Immunocompromised host<sup>1</sup>, n (%) | 4 (19.0) | 15 (16.0) | 0.73 |
| Diabetes mellitus, n (%) | 2 (9.5) | 15 (16.0) | 0.45 |
| Malignancy, n (%) | 0 (0.0) | 8 (8.5) | 0.17 |
| Autoimmune disorder, n (%) | 4 (19.0) | 11 (11.7) | 0.37 |
| Renal disease, n (%) | 0 (0.0) | 15 (16.0) | <0.05 |
| Cardiovascular disease, n (%) | 4 (19.0) | 10 (10.6) | 0.29 |
| Other comorbidities | 3 (14.3) | 15 (16.0) | 0.85 |
| **HLH-2004 criteria<sup>2</sup>** |  |  |  |
| Fever, n (%) | 20 (95.2) | 91 (96.8) | 0.72 |
| Splenomegaly<sup>3</sup>, n (%) | 9 (42.9) | 57 (60.6) | 0.14 |
| Bicytopenia or pancytopenia | 19 (90.5) | 83 (88.3) | 0.78 |
| Hypertriglyceridemia and/or hypofibrinogenemia<sup>4</sup>, n (%) | 9 (42.9) | 51 (54.3) | 0.34 |
| Hemophagocytosis<sup>5</sup>, n (%) | 19 (90.5) | 87 (92.6) | 0.75 |
| Hyperferritinemia<sup>6</sup>, n (%) | 16 (76.2) | 66 (70.2) | 0.58 |
| HLH-2004 score $\geq 5$<sup>7</sup> | 13 (61.9) | 63 (67.0) | 0.65 |

**Abbreviations:** ATT antituberculosis treatment; HLH hemophagocytic lymphohistiocytosis.
Legend: For categorical variables, p-values were calculated using chi-square tests, differences in age was assessed using the Mann-Whitney U-test.
<sup>1</sup>all cases were immunocompromised due to the use of immunomodulatory medication. 4/4 patients on ATT at HLH onset received immunomodulatory medication due to an underlying autoimmune disorder. Of the patients not on ATT at HLH onset receiving immunomodulatory medication, 8/15 had an underlying autoimmune disorder and 7/15 received immunomodulatory medication related to the presence of a transplant or malignancy. Steroids and TNF-inhibitors were the most commonly used immunomodulatory medication; <sup>2</sup>NK-cell activity and soluble CD25 were only measured in 6 and 13 respectively, and therefore not mentioned in the table; <sup>3</sup>splenomegaly was assessed clinically or via imaging; <sup>4</sup>triglycerides $\geq 265$ mg/dL or reported as hypertriglyceridemia and/or fibrinogen $\leq 1.5$ g/L or reported as hypofibrinogenemia; <sup>5</sup>hemophagocytosis in bone marrow, spleen, and or lymph nodes; <sup>6</sup>ferritin $\geq 500$ μg/L or reported as hyperferritinemia; <sup>7</sup>HLH-score was calculated based on available data, including NK-cell activity and soluble CD25 levels.

Pulmonary disease was present in 90.5% (19/21) in TB-HLH patients with a known TB diagnosis, but only in 59.6% (56/96) of patients first presenting with HLH of unknown cause (p = 0.007, **Table 2**). Pulmonary TB followed a miliary pattern in around half of patients in both groups. Most TB-HLH cases showed extrapulmonary TB involvement, with 66.7% (14/21) in those with a confirmed TB diagnosis versus 88.3% (83/94, p = 0.014) presenting with HLH of unknown cause. The distinction between the two groups was even more pronounced when quantifying reported evidence of *M. tuberculosis* in the bone marrow, present in 14.3% (3/21) and 58.5% (55/94) patients respectively (p < 0.001). Notably, this difference was unlikely to be confounded by the frequency of bone marrow testing for *M. tuberculosis* infection as positivity rates of bone marrow investigations for *M. tuberculosis* were lower in patients with a confirmed TB diagnosis vs. those presenting with HLH of unknown cause (30%, 3/10 vs 85.9%, 54/64, p < 0.001, **Figure 3**). T-cell based assays, TST and/or IGRA, conducted in one third of patients, yielded positive results in only 10.5% (4/38) of those tested. Of the IGRA tests performed, 46.7% (7/15) were indeterminate (**Figure 4**).

**Figure 3:**
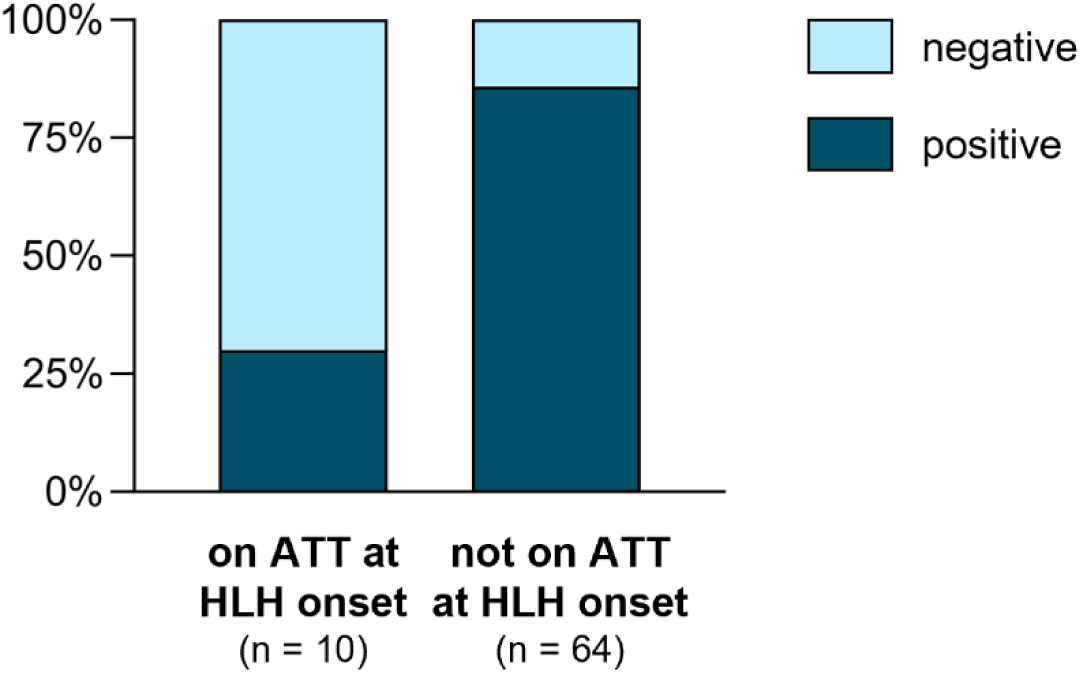
*M. tuberculosis* bone marrow diagnostics by culture, microbiological staining, PCR or histology. Results presented as proportion of all patients in which test bone marrow investigations for evidence of *M. tuberculosis* were conducted. Patients were categorized into patients with known TB developing HLH as a paradoxical reaction while receiving antituberculosis treatment (left); and patients presenting with HLH of unknown cause who were later diagnosed with TB, and who therefore were not receiving antituberculosis treatment at the onset of HLH (right). **Abbreviations: ATT** antituberculosis treatment; **HLH** hemophagocytic lymphohistiocytosis; **PCR** polymerase chain reaction; **TB** tuberculosis.

**Figure 4:**
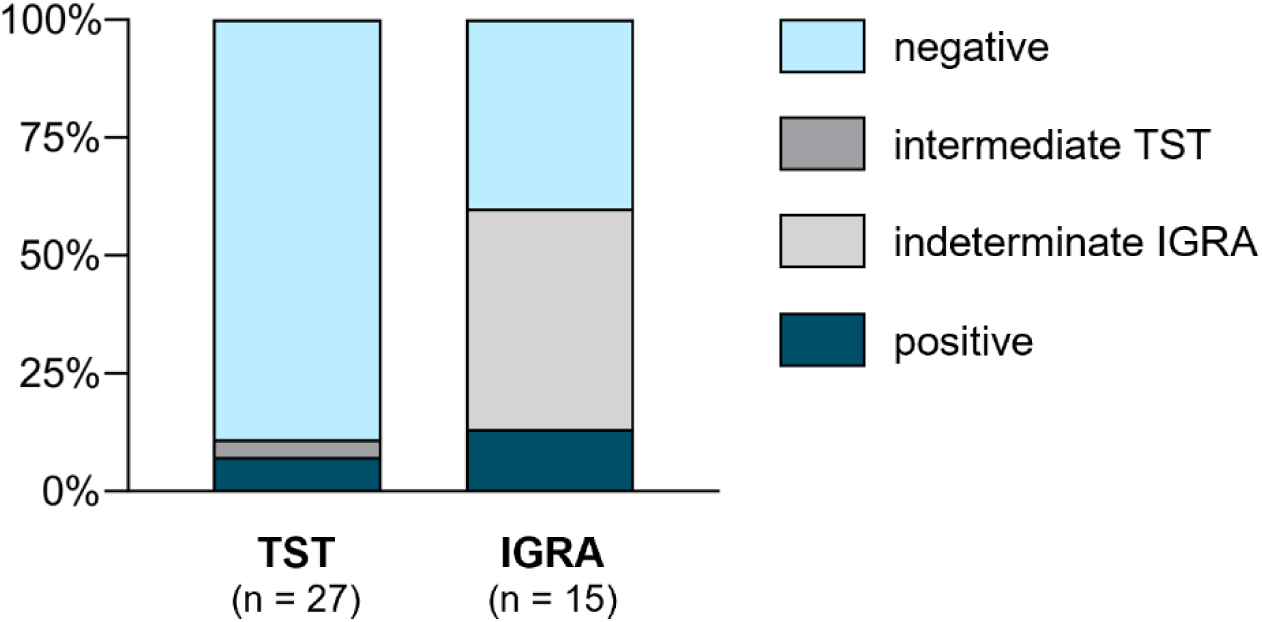
T cell-based immunoassay results in TB-HLH. Tuberculin skin test (left) and interferon gamma release assay (right) test results. Results presented as proportion of all TB-HLH patients in which test results were reported. Four patients had both TST and IGRA results. **Abbreviations: TST** tuberculin skin test; **IGRA** interferon gamma release assay.

**Table 2:** Localization of tuberculosis disease in TB-HLH patients.

|  | On ATT at HLH onset |  |  | Not on ATT at HLH onset |  |  |
| --- | --- | --- | --- | --- | --- | --- |
|  | No evidence of *M. tuberculosis* in bone marrow, n | Evidence of *M. tuberculosis* in bone marrow, n | Total, n (%) | No evidence of *M. tuberculosis* in bone marrow, n | Evidence of *M. tuberculosis* in bone marrow, n | Total, n (%) |
| **No pulmonary involvement, n (%)** |  |  | **2/21 (9.5)** |  |  | **38/94 (40.4)** |
| Solitary lymph node disease, n (%) | 2 | - |  | 3 | - |  |
| Extrapulmonary disease, n (%) | 0 | 0 |  | 15 | 9 |  |
| Solitary bone marrow disease, n (%) | - | 0 |  | - | 11 |  |
| **Pulmonary involvement, n (%)** |  |  | **19/21 (90.5)** |  |  | **56/94 (59.6)** |
| Parenchymal disease, n (%) | 2 | 2 |  | 7 | 7 |  |
| Parenchymal disease with extrapulmonary localization, n (%) | 4 | 0 |  | 5 | 11 |  |
| Miliary pattern, n (%) | 5 | 0 |  | 4 | 12 |  |
| Miliary pattern with extrapulmonary localization, n (%) | 5 | 1 |  | 5 | 5 |  |
| **Total, n (%)** | **18/21 (85.7%)** | **3/21 (14.3%)** | **21** | **39/94 (41.5)** | **55/94 (58.5)** | **94** |

**Abbreviations:** ATT antituberculosis treatment; HLH hemophagocytic lymphohistiocytosis.
Legend: for 11 patients on ATT at HLH onset and 21 patients not on ATT at HLH onset, bone marrow investigations for the evidence of *M. tuberculosis* were not reported. These patients were reported as 'no evidence of *M. tuberculosis* in bone marrow' in this table.

At the time of HLH diagnosis, antituberculosis treatment had already been started in all 21 patients with a known TB diagnosis, while only 80.9% (76/94) of TB-HLH presenting with HLH of unknown etiology received antituberculosis treatment at some point during their hospital stay. Corticosteroids were initiated in 65.2% (75/115). In hospital mortality was 40.0%, and did not significantly differ between those with and without known TB at time of HLH diagnosis (23.8% vs 43.6%, p = 0.094). Older age, presence of comorbidities, hepatomegaly, evidence of *M. tuberculosis* in the bone marrow, no steroid therapy, and not receiving antituberculosis treatment during the disease episode were individually associated with in-hospital mortality. All (18/18) patients not on antituberculosis treatment died, while in hospital mortality was only 27.8% (28/97) in patients started on antituberculosis treatment (p < 0.001). Time from hospital admission to death was 13 days (IQR 7-22) in patients who did not receive antituberculosis treatment (reported for 14/18 patients) and 31 days (IQR 13-42, p = 0.007) who did receive antituberculosis treatment (reported for 22/28 patients). Additionally, in 12/18 patients who did not receive antituberculosis treatment, the diagnosis of tuberculosis was made posthumously.

The multivariable regression analysis demonstrated a trend towards the presence of comorbidities and *M. tuberculosis* in the bone marrow to be independently associated with increased mortality in TB- HLH (Table 3). 100% mortality in all patients not receiving antituberculosis treatment generated a Hauck-Donner effect [25] for TB treatment as a variable in multivariate analyses, precluding accurate estimate of the beta-coefficient and its associated p value.

**Table 3:**
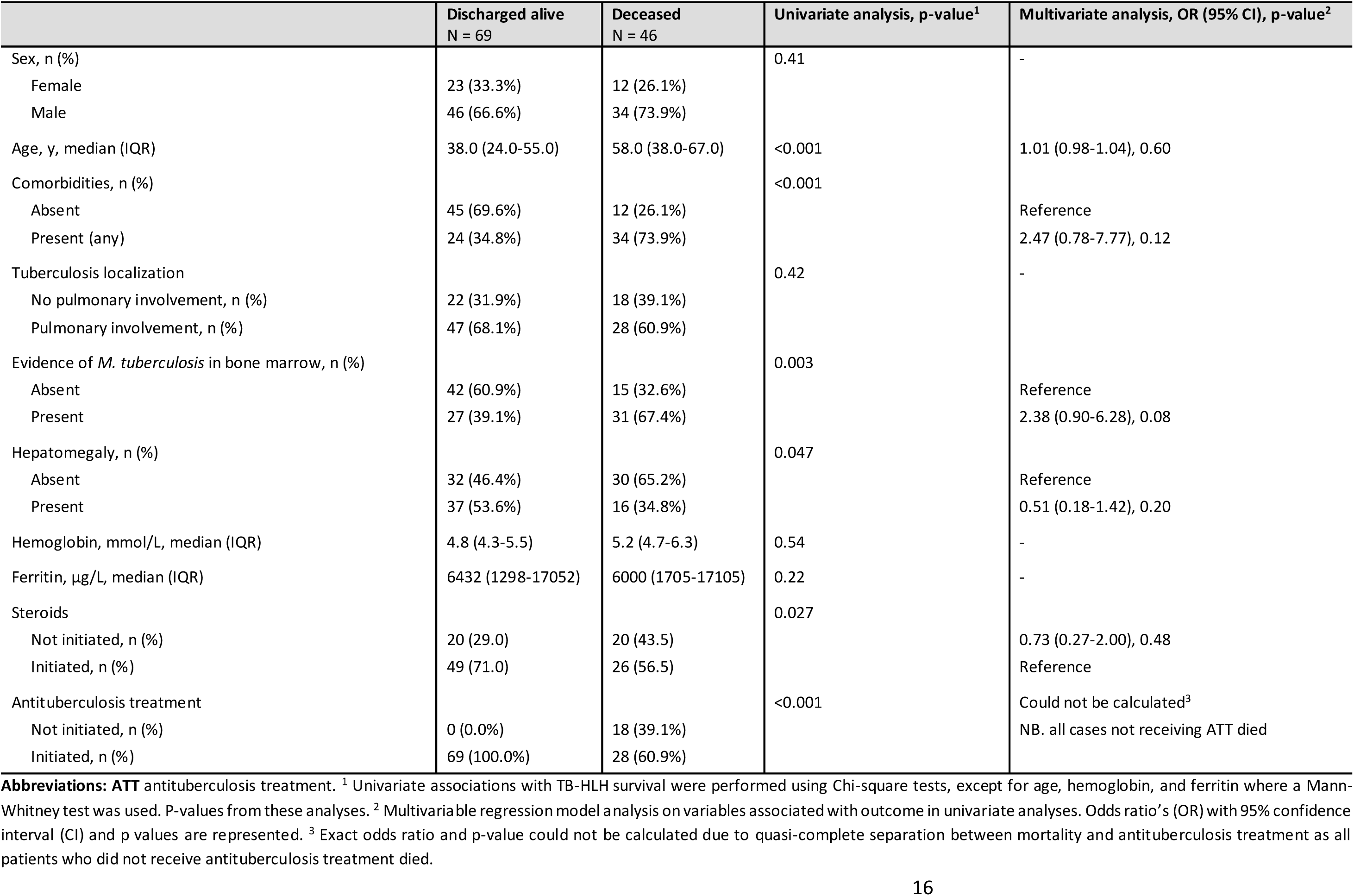
Disease outcomes.

|  | Discharged alive<br>N = 69 | Deceased<br>N = 46 | Univariate analysis, p-value<sup>1</sup> | Multivariate analysis, OR (95% CI), p-value<sup>2</sup> |
| --- | --- | --- | --- | --- |
| Sex, n (%) |  |  | 0.41 | - |
| Female | 23 (33.3%) | 12 (26.1%) |  |  |
| Male | 46 (66.6%) | 34 (73.9%) |  |  |
| Age, y, median (IQR) | 38.0 (24.0-55.0) | 58.0 (38.0-67.0) | <0.001 | 1.01 (0.98-1.04), 0.60 |
| Comorbidities, n (%) |  |  | <0.001 |  |
| Absent | 45 (69.6%) | 12 (26.1%) |  | Reference |
| Present (any) | 24 (34.8%) | 34 (73.9%) |  | 2.47 (0.78-7.77), 0.12 |
| Tuberculosis localization |  |  | 0.42 | - |
| No pulmonary involvement, n (%) | 22 (31.9%) | 18 (39.1%) |  |  |
| Pulmonary involvement, n (%) | 47 (68.1%) | 28 (60.9%) |  |  |
| Evidence of *M. tuberculosis* in bone marrow, n (%) |  |  | 0.003 |  |
| Absent | 42 (60.9%) | 15 (32.6%) |  | Reference |
| Present | 27 (39.1%) | 31 (67.4%) |  | 2.38 (0.90-6.28), 0.08 |
| Hepatomegaly, n (%) |  |  | 0.047 |  |
| Absent | 32 (46.4%) | 30 (65.2%) |  | Reference |
| Present | 37 (53.6%) | 16 (34.8%) |  | 0.51 (0.18-1.42), 0.20 |
| Hemoglobin, mmol/L, median (IQR) | 4.8 (4.3-5.5) | 5.2 (4.7-6.3) | 0.54 | - |
| Ferritin, µg/L, median (IQR) | 6432 (1298-17052) | 6000 (1705-17105) | 0.22 | - |
| Steroids |  |  | 0.027 |  |
| Not initiated, n (%) | 20 (29.0) | 20 (43.5) |  | 0.73 (0.27-2.00), 0.48 |
| Initiated, n (%) | 49 (71.0) | 26 (56.5) |  | Reference |
| Antituberculosis treatment |  |  | <0.001 | Could not be calculated<sup>3</sup> |
| Not initiated, n (%) | 0 (0.0%) | 18 (39.1%) |  | NB. all cases not receiving ATT died |
| Initiated, n (%) | 69 (100.0%) | 28 (60.9%) |  |  |

**Abbreviations:** ATT antituberculosis treatment. <sup>1</sup> Univariate associations with TB-HLH survival were performed using Chi-square tests, except for age, hemoglobin, and ferritin where a Mann-Whitney test was used. P-values from these analyses. <sup>2</sup> Multivariable regression model analysis on variables associated with outcome in univariate analyses. Odds ratio's (OR) with 95% confidence interval (CI) and p values are represented. <sup>3</sup> Exact odds ratio and p-value could not be calculated due to quasi-complete separation between mortality and antituberculosis treatment as all patients who did not receive antituberculosis treatment died.

## Discussion

In this patient-level meta-analysis, we identified 115 adult HIV-negative individuals with TB-HLH. Compared to the general TB population, a much larger proportion of TB-HLH patients show disseminated disease, manifested by extrapulmonary TB, miliary lung disease, or bone marrow involvement. T-cell based assays mostly yielded negative, anergic results, and commencement of antituberculosis treatment was associated with a lower risk of death. Overall, our findings emphasize the importance of considering TB as a cause of HLH, using appropriate diagnostics, and the prompt initiation of TB therapy.

HLH is associated with high mortality [4], and we found TB-HLH to be no different. In the reported cases we identified, 40% of patients died. Importantly, starting treatment for underlying TB disease was the only modifiable factor associated with lower mortality. This supports guidance to identify and treat underlying infectious drivers of secondary HLH, as recommended for viruses (EBV or CMV) and parasites (*Leishmania* spp.)[5]. This also underlines the importance of diagnosing TB-HLH early. Based on the geographical distribution of published TB-HLH cases, we infer there are likely many undiagnosed cases of TB-HLH in high TB-burden countries.

Extrapulmonary TB, particularly bone marrow involvement, was reported in the majority of TB-HLH cases, whereas extrapulmonary TB is less frequent in non-HLH TB case series [26–28]. TB should therefore be considered in patients with unexplained HLH, and the diagnostic workup for TB in suspected or confirmed HLH should include bone marrow histology and microbiological testing for *M. tuberculosis.* If bone marrow sampling is not possible, mycobacterial blood cultures may provide additional diagnostic benefit, but their slow turnaround time means molecular and antigen tests that detect *M tuberculosis*, currently limited to research, may become particularly useful in this setting [29,30].

When TB is diagnosed first, diagnosing HLH can be challenging, as TB can lead to fever, cytopenia and other characteristics of HLH [27]. Hyperferritinemia is often considered specific for HLH, but this has been refuted for adults with HLH [31,32] and we corroborated that a ferritin < 2.000 µg/L or even a normal ferritin does not exclude TB-HLH. In clinical practice, soluble IL2 receptor concentration and especially NK cell activity are not routinely measured, which was also demonstrated in the identified cases. Still, the large majority of patients met ≥5 HLH-criteria Of note, while anemia is most almost universally seen in TB, even in miliary TB it is rare to see leukopenia (< 20%) or thrombocytopenia (<25%)[33]. The high proportion of patients (> 85%) with at least bicytopenia, together with hemophagocytosis in > 90% of patients are the best attainable proof of a high likelihood of true HLH in these patients. Our data suggest that an unusual severe presentation of TB, particularly when disseminated, or unexplained clinical deterioration during antituberculosis treatment, warrants further testing for HLH.

We found a strikingly high incidence of anergy, defined by the absence of IFNy-mediated T cell memory immune responses. A lack of responses to *M. tuberculosis* antigens [34], and mitogen stimulation [35] are associated with miliary TB and in keeping with this, we found very few cases of TB-HLH contained to lungs or lymph nodes [36] Defective adaptive immune responses to *M. tuberculosis* may facilitate persistence and dissemination of *M. tuberculosis* to the bone marrow, a scenario that may further impair protective host defense [37]. Central to HLH pathophysiology is a positive feedback loop between pro-inflammatory cytokines produced by macrophages, with overstimulation but impaired lysis by lymphocytes [2], evidenced by defective NK and CD8+ T-cell cytolytic function in influenza- and dengue-associated HLH [38,39]. The exact pathophysiology of TB-HLH needs further investigation, but our data indicates that removal of mycobacterial antigen drivers through initiation of antituberculosis treatment may be a key step in attenuating pathological hyperinflammation in TB-HLH.

Our study is the first PRISMA-compliant, individual patient level meta-analysis, bringing together all reported adult HIV-negative TB-HLH patients. We overlap 73 adult cases but have added an additional 42 adult cases to those in the most recent review that comprised 116 cases, including 43 that we excluded because they were pediatric, HIV-positive or had missing data [7]. This allowed a priori specified subgroup comparisons and multivariate outcome analyses. Limitations include publication bias and geographical selection inherent to all series of case reports. Further limitations include heterogeneity and selectivity in outcome reporting, and the lack of non-TB secondary HLH groups for comparison. Genetic testing for mutations associated with primary HLH was performed in very few cases, and assessments of immune function were limited to TSTs and IGRAs. For indeterminate IGRA results, individual tube results were lacking, precluding separation of global anergy (negative mitogen tube) versus systemically circulating IFNg yielding nil values above the quality control, which can occur in non-TB HLH [40] and TB-HLH [41]. Lymphocyte counts were available too infrequently to explore their effects on negative or indeterminate IGRA results. More detailed immunophenotyping including quantifying interferon gamma signaling activity and genetic testing is necessary to explore possible primary defects in macrophage or lymphocyte function, while assessing inflammasome function could better inform the pathophysiology underlying the observed hyperinflammation. The limited availability of IGRA results prevented testing for an association between indeterminate IGRA and mortality as has been demonstrated in non-HLH TB [42]. Focusing on HIV-negative individuals prevented us extrapolating our conclusions to the setting of HIV co-infection, in which antiretroviral-associated IRIS reactions share features with HLH [41,43]. Finally, our inference on the importance of prompt antituberculosis treatment commencement in TB-HLH could be prone to survivor bias, and in the absence of data, no conclusions can be made regarding the role of other HLH-targeting therapies used in TB- HLH including corticosteroids.

## Conclusions

Our meta-analysis shows that TB should be considered and bone marrow tested for *M. tuberculosis* in cases of HLH without a known driver, irrespective of patients’ geographical origin or presentation. Moreover, our findings suggest that HLH should be considered in TB patients developing clinical features of hyperinflammation. Mortality in TB-HLH is high and a key modifiable factor in improving survival is prompt initiation of antituberculosis treatment.

## Data Availability

All data produced in the present work are contained in the manuscript.

## Notes

### Acknowledgements

The authors thank Kirsten van Abeelen for statistical advice and help with data visualization.

### Author contribution

LK, GP and AvL conceptualized and designed the study. LK, GP and AvL screened the abstracts for relevance. LK and TS acquired primary data out of the included papers, reviewed for accuracy by SvD, RvC, GP and AvL. LK analyzed the data, followed by interpretation of the data by all authors. LK and TS drafted the article, which was reviewed and approved by all authors.

### Financial support

GP’s time was funded by the UCLH NIHR Biomedical Research Centre. AvL was supported by a Clinical Fellowship of The Netherlands Organization for Health Research and Development (ZonMw, 9032212110006).

### Potential conflicts of interests

None.

## Notes

### Competing Interest Statement

The authors have declared no competing interest.

### Funding Statement

GP time was funded by the UCLH NIHR Biomedical Research Centre. AvL was supported by a Clinical Fellowship of The Netherlands Organization for Health Research and Development - ZonMw, 9032212110006

### Author Declarations

All data used for this systematic review and meta-analysis were derived from openly available, published case reports and series. We defined and followed a pre-specified search strategy according to PRISMA guidelines that was registered with the international prospective register of systematic reviews (PROSPERO, CRD42022349077). The studies that were included in our analyses, and references to their corresponding primary publications, are provided in the supplementary file.

