## Supplementary material for "Tuberculosis-associated hemophagocytic lymphohistiocytosis: diagnostic challenges and determinants of outcome"

**Supplementary table 1:** search strategy

| Database | Coverage Range | Search Executed | Search strategy | # of results |
|---|---|---|---|---|
| PubMed | 1984 to present | October 23rd 2022 | ("Tuberculosis"[Title/Abstract] OR "Tuberculoses"[Title/Abstract] OR "Tuberculous"[Title/Abstract] OR "Tuberculosis"[MeSH Terms] OR "Mycobacterium tuberculosis"[MeSH Terms]) AND ("Hemophagocytic lymphohistiocytosis"[Title/Abstract] OR "Haemophagocytic lymphohistiocytosis"[Title/Abstract] OR "HLH"[Title/Abstract] OR "Hemophagocytic syndrome"[Title/Abstract] OR "Haemophagocytic syndrome"[Title/Abstract] OR "macrophage activation syndrome*"[Title/Abstract]) | 211 records |
| Embase | 1974 to present | October 23rd 2022 | (Tuberculosis.ti,ab,kf. OR Tuberculoses.ti,ab,kf. OR Tuberculous.ti,ab,kf. OR Tuberculosis/ OR Mycobacterium tuberculosis/) AND ((Hemophagocytic lymphohistiocytosis.ti,ab,kf. OR Haemophagocytic lymphohistiocytosis.ti,ab,kf. OR HLH.ti,ab,kf. OR Hemophagocytic syndrome.ti,ab,kf. OR Haemophagocytic syndrome.ti,ab,kf.) OR macrophage activation syndrome*.ti,ab,kf.) | 404 records |
| Global Index Medicus | 1999 to present | October 23rd 2022 | ("Tuberculosis" OR "Tuberculoses" OR "Tuberculous" OR "Tuberculosis" OR "Mycobacterium tuberculosis") AND ("Hemophagocytic lymphohistiocytosis" OR "Haemophagocytic lymphohistiocytosis" OR "HLH" OR "Hemophagocytic syndrome" OR "Haemophagocytic syndrome" OR "macrophage activation syndrome*") | 26 records |

**Supplementary table 2:** variable list

| Data category | Detailed description |
|---|---|
| 1. Record characteristics | a. year of publication;<br>b. geographic region of the reporting authors:<br>c. number of described patients. |
| 2. Patient characteristics | a. Demographics: sex, age;<br>b. Comorbidities: diabetes mellitus, malignancies, autoimmune disorders, coinfections, EBV-status, HIV-status, other comorbidities. |
| 3. Tuberculosis characteristics | a. Disease localization: site of disease, evidence of *Mycobacterium tuberculosis* in the bone marrow;<br>b. Microbiological assessment: species, antibiotic resistance;<br>c. Diagnostics: method of diagnosis, specimen used for diagnostics, time to diagnosis;<br>d. Treatment: time to start treatment, treatment regimen, treatment side effects. |
| 4. Hemophagocytic lymphohistiocytosis characteristics | a. HLH-2004 criteria: fever, body temperature, hepatosplenomegaly, haemoglobin , neutrophil count, ferritin, triglycerides, fibrinogen, soluble CD25, NK-cell activity, evidence of hemophagocytic lymphohistiocytosis;<br>b. H-score;<br>c. Laboratory values: white blood cell count, lymphocyte count, C-reactive protein, lactate dehydrogenase, liver transaminases, γ-glutamyltransferase, alkaline phosphatase, total bilirubin, albumin;<br>d. Time to diagnosis;<br>e. Treatment: time to start treatment, treatment regimen. |
| 5. Immunological assessment | a. Tuberculin skin test: during acute phase of HLH, after recovery;<br>b. Interferon gamma release assay: during acute phase of HLH, after recovery. |
| 6. Patient outcome | a. Complications;<br>b. Mortality;<br>c. Time of follow-up;<br>d. Recurrence of HLH/MAS. |

**Abbreviations:** **HLH** hemophagocytic lymphohistiocytosis; **NK-cell** natural killer cell; **MAS** macrophage activation syndrome

**Supplementary table 3:** Joanna Briggs Institute checklist scores of the case reports included in this review (n = 106)

| Author [ref] | Item number and score | | | | | | | |
|---|---|---|---|---|---|---|---|---|
| | 1 | 2 | 3 | 4 | 5 | 6 | 7 | 8 |
| Aggarwal [1] | Y | Y | Y | Y | Y | Y | Y | Y |
| Al-Mashdali [2] | Y | Y | Y | Y | Y | Y | Y | Y |
| André [3] | Y | Y | Y | Y | Y | Y | U | N |
| Arfa [4] | Y | Y | Y | Y | Y | Y | U | Y |
| Asaji [5] | Y | Y | Y | Y | Y | Y | Y | Y |
| Au [6] | Y | Y | Y | Y | Y | Y | U | N |
| Avasthi [7] | Y | Y | Y | Y | Y | Y | U | N |
| Azzeddine [8] | Y | Y | Y | Y | Y | Y | U | Y |
| Balkis [9] | Y | Y | Y | Y | Y | Y | Y | Y |
| Barnes [10] | Y | Y | Y | Y | Y | Y | Y | N |
| Basu [11] | Y | Y | Y | Y | Y | Y | U | Y |
| Becker [12] | Y | Y | Y | Y | Y | Y | Y | Y |
| Bizid [13] | Y | Y | Y | Y | Y | Y | U | Y |
| Brastianos [14] | Y | Y | Y | Y | Y | Y | U | Y |
| Browett [15] | Y | Y | Y | Y | Y | Y | Y | N |
| Calleja [16] | Y | Y | Y | Y | Y | Y | U | N |
| Campo [17] | Y | Y | Y | Y | Y | Y | U | Y |
| Campos [18] | Y | Y | Y | Y | Y | Y | U | Y |
| Cassim [19] | Y | Y | Y | Y | Y | Y | Y | N |
| Castellano [20] | Y | Y | Y | Y | Y | Y | U | Y |
| Cerme [21] | Y | Y | Y | Y | Y | Y | U | N |
| Chen [22] | Y | Y | Y | Y | Y | Y | U | Y |
| Chen [23] | Y | Y | Y | Y | Y | Y | Y | Y |
| Cherif [24] | Y | Y | Y | U | Y | Y | U | Y |
| Chien [25] | Y | Y | Y | Y | Y | Y | U | Y |
| Chokshi [26] | Y | Y | Y | Y | Y | Y | U | U |
| Claessens [27] | Y | Y | Y | Y | Y | Y | Y | Y |
| Costa [28] | Y | Y | Y | Y | Y | Y | U | N |
| Dalugama [29] | Y | Y | Y | Y | Y | Y | Y | N |
| Dassi [30] | Y | Y | Y | Y | Y | Y | U | Y |
| De Sousa Arantes Ferreira [31] | Y | Y | Y | Y | Y | Y | U | Y |
| Dhawale [32] | Y | Y | Y | Y | Y | Y | U | N |
| Elhence [33] | Y | Y | Y | Y | Y | Y | U | Y |
| Eliopoulos [34] | Y | Y | Y | Y | Y | Y | U | Y |
| Fernandez [35] | Y | Y | Y | Y | Y | Y | U | Y |
| Francois [36] | Y | Y | Y | Y | Y | Y | U | Y |
| Gandotra [37] | Y | Y | Y | Y | Y | Y | U | N |
| Geerdes-Fenge [38] | Y | Y | Y | Y | Y | Y | Y | N |
| Gupta [39] | Y | Y | Y | Y | Y | Y | U | Y |
| Halabi [40] | Y | Y | Y | Y | Y | Y | U | N |
| Hansen [41] | Y | Y | Y | N | Y | Y | U | N |
| Haque [42] | Y | Y | Y | Y | Y | Y | U | Y |
| Hashmi [43] | Y | Y | Y | Y | Y | Y | U | Y |
| Hernandez [44] | Y | Y | Y | Y | Y | Y | U | U |
| Ho [45] | Y | Y | Y | Y | Y | Y | U | N |
| Hui [46] | Y | Y | Y | Y | Y | Y | Y | Y |
| Jain [47] | Y | Y | Y | N | Y | Y | Y | Y |
| Kargupta [48] | Y | Y | Y | Y | Y | Y | U | Y |
| Kessler [49] | Y | Y | Y | Y | Y | Y | U | N |
| Khan [50] | Y | Y | Y | Y | Y | Y | U | Y |
| Ko [51] | Y | Y | Y | Y | Y | Y | U | Y |
| Kursat [52] | Y | Y | Y | U | Y | Y | U | Y |
| Lam [53] | Y | Y | Y | Y | Y | Y | U | N |
| Laxminarayana [54] | Y | Y | Y | Y | Y | Y | U | Y |
| Lee [55] | Y | Y | Y | Y | Y | Y | U | Y |
| Leibowitz [56] | Y | Y | Y | Y | Y | Y | U | N |
| Leon-Sanchez [57] | Y | Y | Y | U | Y | Y | U | Y |
| Lombardo [58] | Y | Y | Y | Y | Y | Y | U | Y |

| | | | | | | | | |
|---|---|---|---|---|---|---|---|---|
| Long [59] | Y | Y | Y | Y | Y | Y | U | N |
| Maheswar [60] | Y | Y | Y | Y | Y | Y | U | U |
| Manski [61] | Y | Y | Y | Y | Y | Y | U | Y |
| Matsuura-Otsuki [62] | Y | Y | Y | Y | Y | Y | U | N |
| Mbivzo [63] | Y | Y | Y | Y | Y | Y | U | N |
| Naha [64] | Y | Y | Y | Y | Y | Y | U | Y |
| Nakamura [65] | Y | Y | Y | Y | Y | Y | U | N |
| Ohata [66] | Y | Y | Y | Y | Y | Y | U | Y |
| Padhi [67] | Y | Y | Y | Y | Y | Y | U | Y |
| Padhi [68] | Y | Y | Y | Y | Y | Y | U | N |
| Poornachandra [69] | Y | Y | Y | Y | Y | Y | Y | N |
| Quiquandon [70] | Y | Y | Y | Y | Y | Y | U | Y |
| Rakotoson [71] | Y | Y | Y | Y | Y | Y | U | Y |
| Ranjan [72] | Y | Y | Y | U | Y | Y | U | Y |
| Rao [73] | Y | Y | Y | Y | Y | Y | U | N |
| Rathnayake [74] | Y | Y | Y | U | Y | Y | U | Y |
| Roca [75] | Y | Y | Y | Y | Y | Y | U | N |
| Rodriguez [76] | Y | Y | Y | Y | Y | Y | U | Y |
| Rodriguez-Medina [77] | Y | Y | Y | Y | Y | Y | U | Y |
| Rosales-Castollo [78] | Y | Y | Y | Y | Y | Y | U | Y |
| Ruiz-Argüelles [79] | Y | Y | Y | Y | Y | Y | Y | N |
| Saez-Conzalez [80] | Y | Y | Y | Y | Y | Y | U | N |
| Sandrini [81] | Y | Y | Y | Y | Y | Y | U | N |
| Satomi [82] | Y | Y | Y | Y | Y | Y | U | N |
| Sayes [83] | Y | Y | Y | Y | Y | Y | U | Y |
| Schippers [84] | Y | Y | Y | Y | Y | Y | U | Y |
| Seminari [85] | Y | Y | Y | Y | Y | Y | U | N |
| Sharma [86] | Y | Y | Y | Y | Y | Y | U | Y |
| Shea [87] | Y | Y | Y | Y | Y | Y | U | Y |
| Shi [88] | Y | Y | Y | Y | Y | Y | U | Y |
| Shin [89] | Y | Y | Y | Y | Y | Y | Y | Y |
| Shiu [90] | Y | Y | Y | Y | Y | Y | U | Y |
| Shoureshi [91] | Y | Y | Y | Y | Y | Y | U | Y |
| Singha [92] | Y | Y | Y | Y | Y | Y | U | Y |
| Su [93] | Y | Y | Y | Y | Y | Y | U | Y |
| Subhash [94] | Y | Y | Y | Y | Y | Y | U | Y |
| Takahashi [95] | Y | Y | Y | Y | Y | Y | U | Y |
| Talluri [96] | Y | Y | Y | Y | Y | Y | U | Y |
| Thiam [97] | Y | Y | Y | Y | Y | Y | U | N |
| Troncoso Mariño [98] | Y | Y | Y | Y | Y | Y | U | N |
| Trovik [99] | Y | Y | Y | Y | Y | Y | U | Y |
| Tseng [100] | Y | Y | Y | Y | Y | Y | Y | N |
| Undar [101] | Y | Y | Y | Y | Y | Y | U | Y |
| Vaiphei [102] | Y | Y | Y | Y | Y | Y | U | N |
| Wang [103] | Y | Y | Y | Y | Y | Y | U | N |
| Weintraub [104] | Y | Y | Y | Y | Y | Y | U | N |
| Yang [105] | Y | Y | Y | Y | Y | Y | U | Y |
| Ye [106] | Y | Y | Y | Y | Y | Y | U | N |
| Number of studies applying the item | 106 | 106 | 106 | 99 | 106 | 106 | 18 | 64 |

**Abbreviations: Y** yes; **N** no; **U** unclear; **NA** not applicable.

Items of the JBI checklist: 1) were patient's demographic characteristics clearly described? 2) was the patients history clearly described and presented as a timeline? 3) was the current clinical condition of the patient clearly described? 4) were diagnostic tests or assessment methods and the results clearly described? 5) was the intervention or treatment procedure(s) clearly described? 6) was the post intervention clinical condition clearly described? 7) were adverse events (harms) or unanticipated events identified and described? 8) Does the case report provide takeaway lessons.

**Supplementary table 4:** Laboratory values at the time of HLH-presentation.

| | N | On ATT at HLH onset | N | Not on ATT at HLH onset | p-value |
|---|---|---|---|---|---|
| Temperature, °C, median (IQR) | 8 | 39.1 (38.4-39.9) | 45 | 39.0 (38.8-39.6) | 0.75 |
| Hemoglobin, mmol/L, median (IQR) | 14 | 4.8 (4.7-6.2) | 71 | 5.1 (4.3-5.5) | 0.81 |
| White blood cell count, x $10^9$/L, median (IQR) | 12 | 2.2 (1.6-3.6) | 72 | 2.6 (1.6-4.2) | 0.76 |
| Lymphocyte count, x $10^9$/L, median (IQR) | 4 | 0.3 (0.1-0.4) | 24 | 0.5 (0.3-0.8) | 0.10 |
| Neutrophil count, x $10^9$/L, median (IQR) | 6 | 1.3 (0.6-2.6) | 28 | 1.0 (0.6-2.6) | 0.60 |
| Thrombocyte count, x $10^9$/L, median (IQR) | 15 | 41.0 (5.0-73.0) | 78 | 47.0 (22.8-90.5) | 0.41 |
| Triglycerides, mg/dL, median (IQR) | 9 | 449.0 (176.6-463.3) | 38 | 314.0 (223.6-380.7) | 0.41 |
| Fibrinogen, g/L, , median (IQR) | 7 | 1.8 (1.4-3.8) | 33 | 1.4 (0.8-3.0) | 0.56 |
| Ferritin, µg/L, median (IQR) | 15 | 6800.0 (3032.0-17209.0) | 51 | 5200.0 (996.0-16854.0) | 0.65 |

**Abbreviations: ATT** antituberculosis treatment; **HLH** hemophagocytic lymphohistiocytosis.

Legend: p-values were calculated using Mann-Withney U-tests.

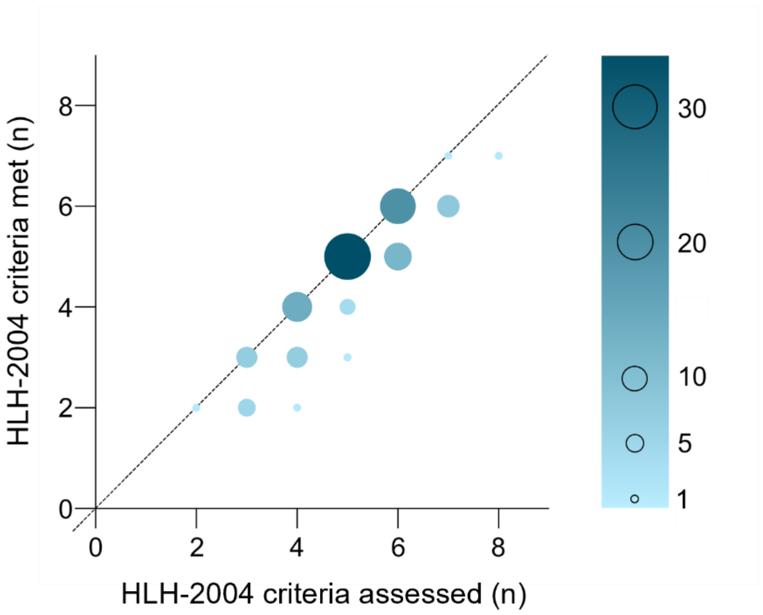


**Supplementary figure 1: HLH-2004 criteria [107].** Number of HLH-2004 criteria assessed (x-axis) plotted against the number of HLH-2004 criteria met (y-axis). The diagonal represents x = y. **Abbreviations: HLH** hemophagocytic lymphohistiocytosis.
